# Passive Sensing of Gait and Medication-related Fluctuations in Parkinson’s Disease

**DOI:** 10.1101/2025.11.12.25340068

**Authors:** Juyoung Jenna Yun, Charalambos Hadjipanayi, Arya Jahangiri, Alan Bannon, Timothy G Constandinou, Shlomi Haar

## Abstract

Gait impairment is a hallmark symptom of Parkinson’s disease (PD). However, traditional clinical assessments cannot capture real-world motor fluctuations, as they are sparsely performed. This study was designed to test and validate the use of nearables and passive sensing technologies, including Kinect RGB-D cameras and ultra-wideband (UWB) radar, for continuous, objective assessment of gait fluctuations in PD within a home-like setting. Fifteen PD patients with mild symptoms and fourteen age-and sex-matched healthy controls (HC) performed 4-meter walking tasks in a living lab facility. Patients repeated the task during both “ON” and “OFF” states of their daily medication cycle. Gait features, including stride length, stride time, and gait speed, were extracted from Kinect, radar, and a ground-truth smart floor. Data were analyzed to evaluate inter-sensor agreements and detect group-level differences. Stride time demonstrated the highest agreement between devices (*r*=0.903), while stride length showed weaker agreement (*r*=0.779), with Kinect tending to overestimate. Despite lower agreement, stride length from both Kinect and radar successfully distinguished PD OFF from HC (camera *q*=0.020; radar *q*=0.005) and radar was able to further differentiate ON and OFF states (*q*=0.020). Neither device differentiated PD ON from HC, indicating medication reduced observable gait differences. This study demonstrates that passive, contact-free sensing technologies such as depth cameras and UWB radars can effectively monitor gait in PD within naturalistic environments. While some spatial metrics, like stride length, show device discrepancies, both systems reliably capture gait patterns and medication-dependent changes, supporting their use for longitudinal, real-world monitoring of Parkinson’s motor symptoms.

## Introduction

Gait disturbances are some of the most common and disabling motor symptoms in Parkinson’s Disease (PD), with freezing of gait affecting up to 80% of individuals with advanced disease(1,2). PD motor symptoms are highly multidimensional, varying considerably across individuals and fluctuating throughout the day in response to medications such as levodopa(3). Currently, the gold standard for assessing disease severity and progression is the Unified Parkinson’s Disease Rating Scale (UPDRS). However, the motor examination part of the UPDRS (known as part III) has several well-recognized limitations, including intra- and inter-rater variability and structured tasks that may not reflect natural movement(4–6). Moreover, the infrequent nature of this measurement during clinical visits limits the sensitivity to the full range of motor fluctuations occurring throughout the day(5).

Marker-based motion capture and instrumented walkways provide a gold standard for quantitative gait analysis(7–9). Yet, those are expensive unscalable setups. In contrast, wearable sensors have emerged as a cost-effective tool for quantifying gait and have become widely used in both clinical and home settings, providing accessible alternatives to traditional laboratory equipment(10–13).

The most common wearables are smartwatches, due to their familiarity and social acceptability, promoting higher compliance for daily use(14,15). Smartwatches can estimate gait parameters using their inertial measurement unit (IMU)(16,17). However, their accuracy can be strongly altered by arm swing patterns in PD patients or reduced arm swing in older adults(16,18,19). Lumbar sensors offer better precision for step detection and temporal metrics and have shown potential in differentiating PD subtypes through gait biomarkers(20,21). However, these wearables are sensitive to accurate placement and consistent orientation on the lumbar spine, where small deviations can distort gait asymmetry and degrade data quality(22,23).

Passive motion tracking technologies are uniquely suited for free-living applications, as they provide uninterrupted and long-term insights into symptom progression without requiring charging, user interaction, or compliance, allowing a truly “install and forget” capability(24,25). RGB-depth cameras like the Microsoft Kinect yield equivalent spatiotemporal gait measurements with comparable accuracy to gold-standard gait assessment methods such as marker-based motion capture (26–29). These systems avoid the need for reflective markers and specialized laboratory setups or dedicated floor surfaces, and thus are practical for home deployments. Despite these benefits, their performance can be influenced by environmental factors such as lighting conditions and occlusions, posing challenges for protocol standardization and measurement consistency(30,31).

Radar systems offer complementary advantages, including enhanced privacy and the ability to detect subtle movements even through clothing, with high temporal resolution to capture gait dynamics that can identify gait disorders or reveal early signs of neurological conditions such as PD(32–35). Our prior work demonstrated that low-power and cost-effective ultra-wideband (UWB) radar can accurately quantify spatiotemporal gait parameters in healthy participants during both normal and asymmetric walking, yet each unit inherently provides a one-dimensional field of view, limiting the accuracy of spatial measurements, unless multiple sensors are combined(35,36).

In this study, we validate the use of Kinect cameras and UWB radars for objective gait assessment in PD within a fully functioning living environment. Importantly, our approach captures natural fluctuations observed in PD patients across the day, including medication “on” effects (ON) and gradual “wearing-off” (OFF) periods, without artificially inducing extreme medication states. This allowed participants to engage in daily activities while continuous, passive motion tracking was used to characterize gait patterns, providing ecologically valid insights that complement standard clinical assessment.

## Methods

### Participants

We recruited 15 individuals with Parkinson’s disease (PD) and 14 age-matched healthy controls (HC). For both PD and HC, exclusion criteria included any neurological disorders or gait impairments that could influence locomotor performance. All participants gave written informed consent before participation in the study. The study was approved by the Imperial College Research Ethics Committee (ICREC) and conducted in accordance with the Declaration of Helsinki.

### Study Procedures

The study was conducted in the Living Lab at Imperial College London(37), a facility designed to replicate a fully functioning home environment with standard domestic furnishings and appliances (Figure 1). For PD participants, each session was scheduled so that patients took their regular medication dose upon arrival, after which preliminary paperwork was completed while waiting for the medication to take effect. Motion data for the ON medication state were collected approximately one hour after medication intake. To capture the “wearing-off” (OFF) state, corresponding motion data were collected immediately before the patients’ next scheduled medication dose, representing the period of maximal symptom re-emergence within their daily cycle. In both ON and OFF states, motor symptom severity was assessed using the UPDRS motor examination (known as part III). In line with the UPDRS gait item, participants performed a straight walk approximating the 10-metre standard. For feasibility within the Living Lab, a 4m path was used, allowing consistent observation of a minimum of four gait cycles per trial. HC participants performed the walking task once and without a full UPDRS motor examination.

**Figure 1.**
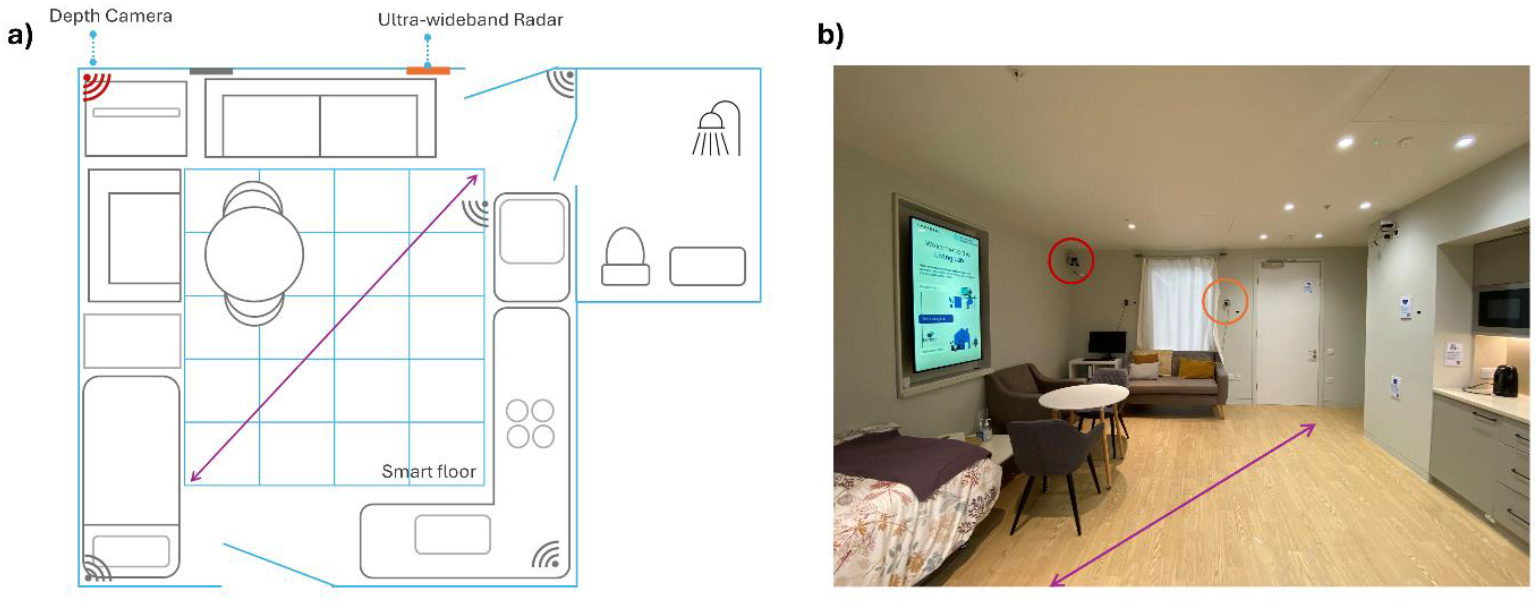
Environmental sensor layout in the Living Lab. **(a)** Schematic of the home-like experimental environment equipped with multimodal sensors. Ultra-wideband radars (rectangle icons – the one in orange was used here) and depth cameras (wave-emission icon – the one in red was used here) were installed around the room. The 4-meter walking pass is marked with a purple arrow. The smart floor system (blue grid) embedded beneath the floor tiles recorded participants’ location while walking. **(b)** Photograph of the Living Lab from the same perspective as panel (a). The radar is highlighted in an orange circle, whereas the camera is in a red circle. The walking trajectory is indicated again with a purple arrow.

During the assessment, three sensing modalities were operated simultaneously, including Microsoft Azure Kinect DK, Novelda XeThru UWB radar, and SensFloor room terminal systems. One Kinect camera and one UWB radar, selected based on their optimized placement relative to the walking path, were used in this study (positioning highlighted in Figure 1). SensFloor data were used as the reference (“ground truth”) for walking speed alignment.

### Camera data

The Azure Kinect DK captured 3D skeletal data from 32 joints at 30Hz. Data were recorded as timestamped JSON files and processed in Python 3.12. The preprocessing pipeline included manual validation of walk trial timestamps, temporal segmentation, and participant identification to exclude any false detections. The three-dimensional joint positions of the pelvis and bilateral ankles were transformed into a walking-aligned coordinate frame based on the pelvis trajectory and floor orientation. Initial contact events were identified from alternating extrema in the filtered anterior–posterior distance between the left and right ankles, from which the step and stride gait parameters were derived.

### Radar data

Radar data were acquired from the sensor positioned closer to the walking path, whose sensing direction was more closely aligned with the walking trajectory and therefore provided higher accuracy when estimating movement along the path length(36). Radar signals were sampled at 500Hz with a distance range resolution of 5.1cm. Signal acquisition, filtering, and post-processing followed the same pipeline as our previously published work(36), implemented in MATLAB (R2024a). This included suppression of reflections from stationary objects and background noise, radar signal demodulation, and data analysis in the range–Doppler domain. From the processed data, relative distance (range) and velocity (Doppler) trajectories of the torso and feet were extracted. Because the walking path was not perfectly aligned with the sensor’s primary viewing direction, a geometric correction was applied to reflect movement along the walking direction. Initial contact events were identified as local minima in the feet velocity trajectories. The distance and timing information of the torso and initial contact events were then used to estimate the selected gait parameters.

### Floor data

SensFloor (Future-Shape GmbH) is a capacitive sensor system. It consists of a 3mm thick textile underlay with built-in sensor electronics. SensFloor measures, through a flooring layer on top of it, changes in the electric capacitances on a triangular grid of sensor fields(38). SensFloor “tracks” position at an approximately 32cm*32cm spatial resolution and a temporal resolution of 100ms. From these data, we derived speed (m/s) by calculating successive x and y coordinates for the same object during the 4m walk. SensFloor provides a continuous approximation of centroids rather than foot contacts. Thus, the torso’s center-of-mass movement is the most physiologically comparable signal, whereas stride or step metrics depend on limb-specific information that the floor does not capture.

### Gait Metrics

Step length was defined as the anterior–posterior distance (in meters) between initial contacts of opposite feet, whereas stride length is the distance covered by the same foot between two successive initial contacts. Individual step lengths are calculated as the distance between consecutive initial contact points, and stride length is computed as the sum of two consecutive step lengths. Stride length is reported as the mean of all valid values across the recording session. Step time is defined as the temporal interval (in seconds) between initial contacts of opposite feet, whereas stride time is the interval between two successive initial contacts of the same foot. Step times are calculated as the time interval between consecutive initial contact points, and stride time is derived as the sum of two consecutive step times. Mean stride time is then computed from all valid strides per participant.

Stride (or gait) speed is defined as the average linear velocity (in meters/second) of a complete stride, derived from detected gait events and reflecting the mean forward walking speed during a gait cycle. It is calculated as stride length divided by stride time, computed separately for each stride and then averaged across all strides in each session. Pelvis or torso speed is defined as the instantaneous linear velocity of the pelvis joint or torso during walking, computed directly from continuous joint trajectories as a kinematic measure of the body’s center movement in the walking direction. It is derived from the time-differentiated displacement of the pelvis joint (Kinect) or torso (radar) in the walking direction, and mean pelvis or torso speed is reported across all valid gait segments for each participant. When SensFloor data are available, torso speed is estimated from the temporal sequence of pressure activations as the center of pressure progresses forward across sensor tiles in the walking direction, allowing calculation of mean forward speed over the gait cycle.

Symmetry index (SI) is defined as the magnitude of gait symmetry between the two legs. This is computed using step length and step time, as these directly reflect inter-limb coordination and asymmetry between sides. Stride metrics, by encompassing both legs, inherently mask such step-level asymmetries. Since radar cannot distinguish between which step originates from which leg (left/right or paretic/non-paretic), the absolute value was taken to make the metric direction-agnostic. Following the definition by Patterson et al.(39), this approach was also applied to camera and UPDRS data, thereby expressing only the magnitude of asymmetry as:

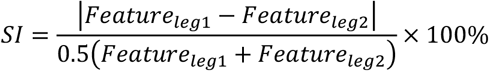

### Statistical Analysis

Statistical analyses were performed using Python. The study employed a cross-sectional design. Descriptive statistics were calculated for movement data obtained from each device, and results for continuous variables are presented as mean ± standard deviation (SD). Prior to parametric analyses, data were tested for normality using the Shapiro–Wilk and Anderson–Darling tests, and for homogeneity of variance using Levene’s and Bartlett’s tests. Independence between the PD and HC groups was maintained by the study design, and paired comparisons were used within the PD group to examine differences between medication states. While using the parametric Pearson’s r, robustness was validated through leave-one-out (LOO) resampling. This approach ensured the stability of the correlation by identifying potential outlier-driven influences. This was further reported with Root Mean Square Error (RMSE) of best fit alongside mean percentage. Paired t-tests were used for within-PD comparisons between ON and OFF states. Statistical significance was defined as p < 0.05. For analyses involving multiple comparisons, q-values were calculated using false discovery rate (FDR) correction.

## Results

A total of 15 individuals with PD and 14 age- and sex-matched HC participants took part in the study (age: *p*=0.346; sex: *p*=0.836) (Table 1a). PD symptoms severity fluctuated across the ON and OFF states of the participants’ medication cycle (Table 1b). A significant difference in the UPDRS motor examination score between states (*p*=0.020) reflects expected dopaminergic improvements in motor symptoms during ON medication. Gait item and symmetry indices derived from the UPDRS were also reported, with no significant differences between states, suggesting the participants’ gait had not changed enough to be captured by the gait measures of the UPDRS.

**Table 1.** Participants’ demographics and clinical characteristics. **(a)** Group comparison between healthy controls (HC) and people with Parkinson’s disease (PD; n = 15). Values represent group means. No significant differences were observed in age, gender distribution, or handedness between groups. Levodopa equivalent daily dose (LEDD) is shown for the PD group. **(b)** Comparison of PD participants’ UPDRS subscores and gait symmetry index between ON and OFF medication conditions.

| a) |  |  |  | b) |  |  |  |
| --- | --- | --- | --- | --- | --- | --- | --- |
|  | HC (n=14) | PD (n=15) | P-value |  | PD ON (n=15) | PD OFF (n=15) | P-value |
| Age | 70.8 | 68.0 | 0.346 | UPDRS III – Gait | 0.53 | 1.07 | 0.055 |
| Gender (male/total) | 10/14 | 10/15 | 0.836 | UPDRS III – Total | 12.7 | 20.5 | **0.020** |
| Handedness (right/left/both) | 10/3/1 | 13/2/0 | 0.637 | Symmetry Index (R-L) | 37.1 | 54.5 | 0.198 |
| LEDD (mg) (min-max) | - | 356 (37.5-850) | - |  |  |  |  |

### Ground-truth validation

Torso speed derived from camera and radar data was compared with floor-measured walking speed (Figure 2a). Both modalities showed strong correlations with the floor measurements, with overall correlation coefficients exceeding 0.90 (*p*<0.0001 for all states). The camera data yielded an overall RMSE of 0.05 m/s (5.25% of the mean) and *r* = 0.922 (HC: *r*=0.942; ON PD: *r*=0.867; OFF PD: *r*=0.959) while radar showed a comparable performance with an overall RMSE of 0.081 m/s (8.45% of the mean) and r=0.900 (HC: *r*=0.841; ON PD: *r*=0.968; OFF PD: *r*=0.874). The strong correlations observed across all groups demonstrate that both sensors capture center-of-mass translation consistently.

**Figure 2.**
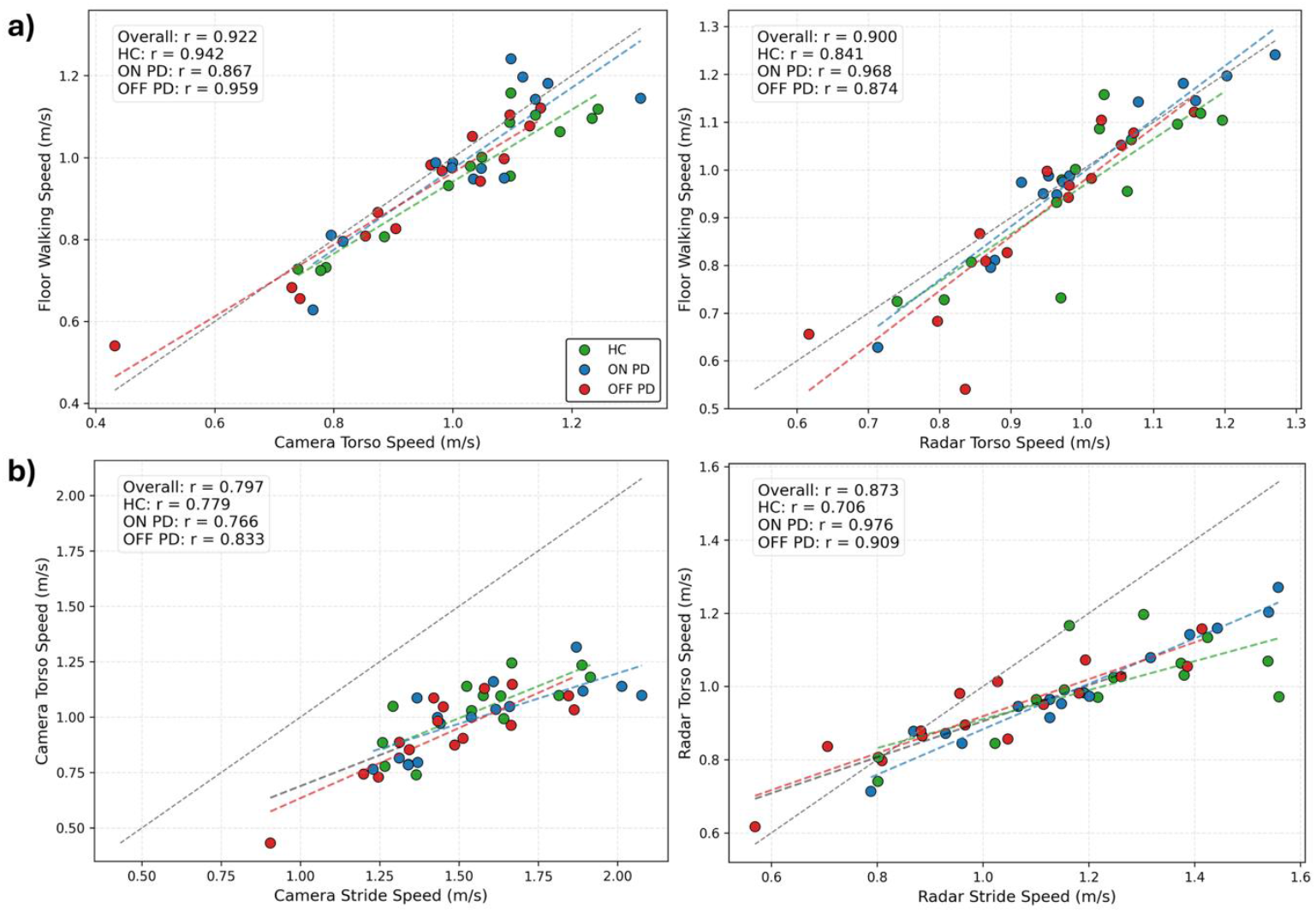
Measurements of walking speed across devices. **(a)** Correlations between smart floor walking speed and camera (left) or radar (right) derived torso speed for healthy controls (HC, green), and Parkinson’s participants (PD) in medication ON (blue), and OFF (red) states. Dashed lines denote linear fits, and the identity line is dotted black. Correlation coefficients (r) are reported for each group and overall. Correlation coefficients (r) are reported for each group and overall. **(b)** Correlations between stride- and torso-level walking speeds derived from the same modality – camera (left) or radar (right). Legends are the same as in (a).

### Agreement within each device

Within-device comparisons were conducted to examine how each sensing modality captures differences between torso speed and foot-derived stride speed (Figure 2b). Camera-derived measures showed an RMSE of 0.104m/s (10.5% of the mean) and overall correlation of *r*=0.797, *p*<0.0001 (HC: *r*=0.779, *p*<0.001; ON PD: *r*=0.766, *p*<0.001; OFF PD: *r*=0.833, *p*<0.0001), while radar demonstrated even better association with an overall RMSE of 0.067m/s (6.88% of the mean) and *r*=0.873, *p*<0.0001 (HC: *r*=0.706, *p*<0.01; ON PD: *r*=0.976, *p*<0.0001; OFF PD: *r*=0.909, *p*<0.0001). The camera data showed a slightly greater variability around the regression line at higher stride speeds, whereas radar measurements were more tightly clustered around the regression line, indicating potentially higher consistency in the velocity estimation. The slope of the regression lines deviating from the identity line in both modalities indicates systematically higher stride speeds compared to torso speeds, reflecting the expected difference between limb-driven forward thrust and center-of-mass translation. This observation highlights the distinction between segmental limb motion and whole-body movement, as torso-based measures primarily capture postural displacement rather than full gait dynamics.

### Agreement between devices

Figure 3 illustrates four key gait-related features, including stride length, stride time, stride speed, and torso speed, compared between camera and radar. Among these, stride time (Figure 3c) showed the strongest inter-device association with RMSE=0.048s (less than 1% of the mean) and *r*=0.903 (*p*<0.0001), demonstrating robust temporal agreement between devices. Stride length (Figure 3a) showed a slightly weaker agreement between devices, with RMSE=0.169m (12.2% of the mean) and *r*=0.779 (*p*<0.0001), where camera measurements tended to overestimate distances relative to radar. Stride speed (Figure 3e), which is a function of stride length and time, displayed a very similar trend with RMSE=0.143m/s (12.55% of mean) and *r*=0.81 (*p*<0.0001). The torso speed (Figure 3g) demonstrated slightly strong agreement between devices with RMSE=0.079m/s (8.10% of the mean) and *r*=0.819 (*p*<0.0001).

**Figure 3.**
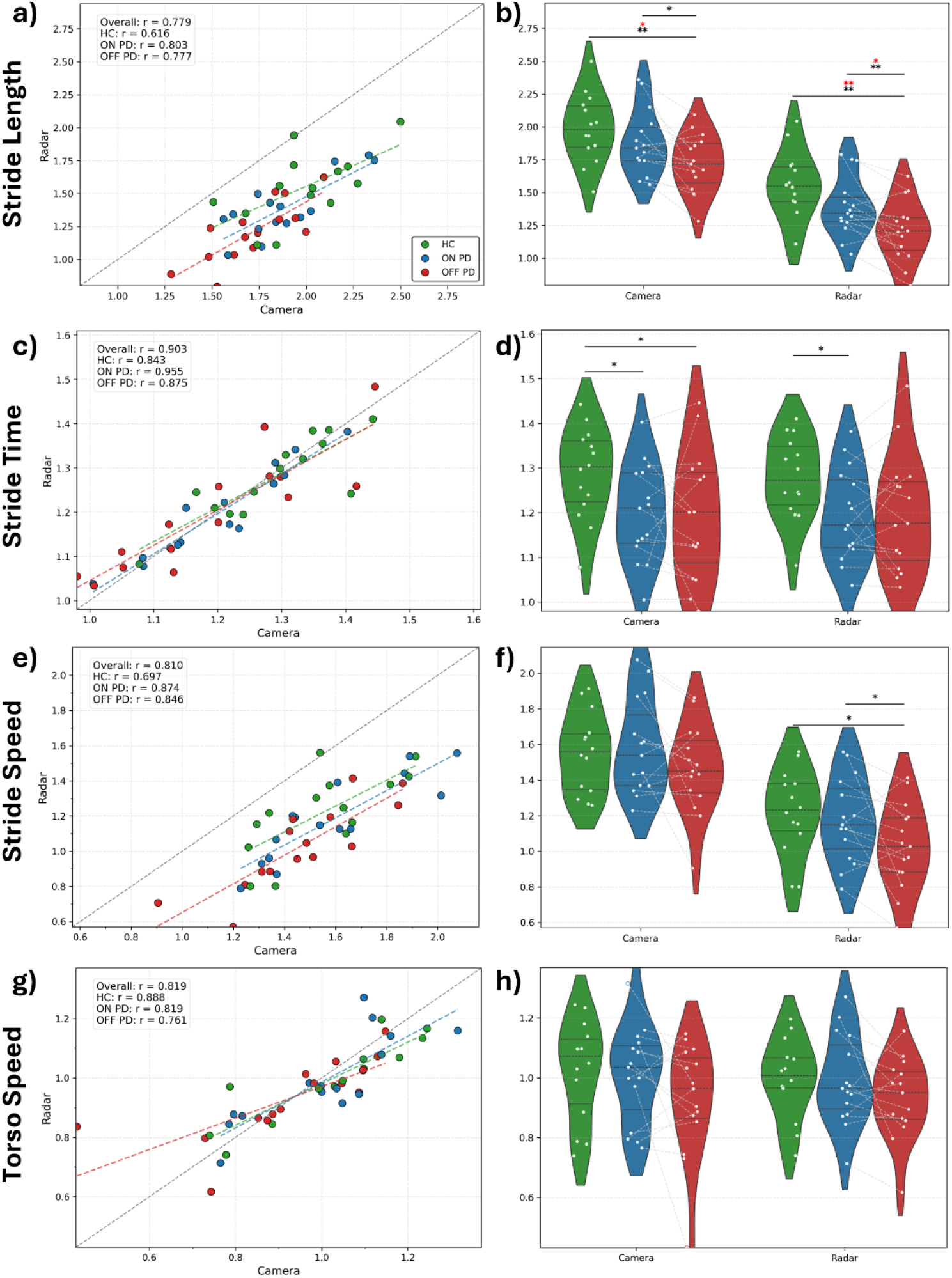
Gait metrics across sensing modalities. Left panels show correlations between camera- and radar-derived metrics: **(a)** stride length, **(b)** stride time, **(c)** stride speed, and **(d)** torso speed. Legends are the same as in Figure 2. Right panels display corresponding violin plots comparing the metrices across the groups, healthy controls (green), and Parkinson’s participants in medication ON (blue), and OFF (red) states. Black asterisks mark uncorrected statistically significant differences (p < 0.05), and red asterisks mark FDR-corrected significant differences (q<0.05). For each asterisk, p/q<0.05 (*), <0.01 (**),<0.001 (***).

### Group and state differences

For stride length (Figure 3b), the radar successfully distinguished differences between HC and PD OFF participants (*q*=0.005) and between PD ON and OFF states (*q*=0.020). Similarly, the camera showed a weaker but still significant difference between HC and PD OFF participants (*q*=0.020) but could not significantly discriminate between ON and OFF states after correction for multiple comparisons (*p*=0.018, *q*=0.070). Neither device differentiated HC from PD ON participants, suggesting that medication substantially reduces stride length differences relative to controls. For stride time (Figure 3d), group-level differences were modest, with camera data showing nominal differences between HC and PD ON (*p*=0.031, *q*=0.128) and between HC and PD OFF (*p*=0.070, *q*=0.099), and radar showing a difference between HC and PD ON (*p*=0.049, *q*=0.124); however, none reached significance after FDR correction. For stride speed (Figure 3f), radar showed nominal differences between HC and PD OFF (*p*=0.037, *q*=0.074) and between PD ON and OFF (*p*=0.027, *q*=0.053), though again these did not survive the FDR correction. Finally, for torso speed (Figure 3h), there were no significant group-dependent differences.

### Symmetry Index Comparison

The relationship between the gait symmetry index and UPDRS motor symmetry index scores was assessed for both camera (Figure 4a) and radar (Figure 4b) data. Camera-derived step length asymmetry showed a moderate positive correlation with UPDRS asymmetry (*r*=0.547, *p*=0.002), with significant associations in both medication states (PD ON: *r*=0.605, *p*= 0.016; PD OFF: *r*=0.558, *p*=0.023). Yet, this was highly variable (overall RMSE=3.85, 72.6% of the mean). Step time asymmetry from the camera also showed a positive association (*r*=0.474, *p*=0.008), which was significant in the ON state (*r*=0.590, *p*=0.021) but not in the OFF state (*r*=0.371, *p*=0.182), yet again, highly variable (overall RMSE=4.17, 88.1% of the mean). The HC step metrics from the camera are shown as 95% reference intervals: [1.38, 5.03] for length asymmetry and [3.33, 6.71] for step time asymmetry, and overlap with the lower end of PD asymmetry, aligning with expectations. In contrast, radar-derived metrics for step length and step time asymmetry demonstrated no significant relationship with UPDRS asymmetry. Step length symmetry from radar was uncorrelated with clinical asymmetry (*r* = –0.008, *p*=0.966; PD ON: *r* = – 0.074, p = 0.794; PD OFF: *r* = –0.026, *p* = 0.930), and step time symmetry exhibited a negligible negative association (*r* = –0.226, *p*=0.230; PD ON: *r* = –0.359, *p* = 0.186; PD OFF: *r* = –0.139, *p* = 0.625). The 95% reference intervals for HC radar data were [35.86, 52.52] for step length asymmetry and [15.30, 23.74] for step time asymmetry, and overlapped with the middle to high end of PD asymmetry, highlighting the invalidity of the asymmetry measures extracted from a single radar.

**Figure 4.**
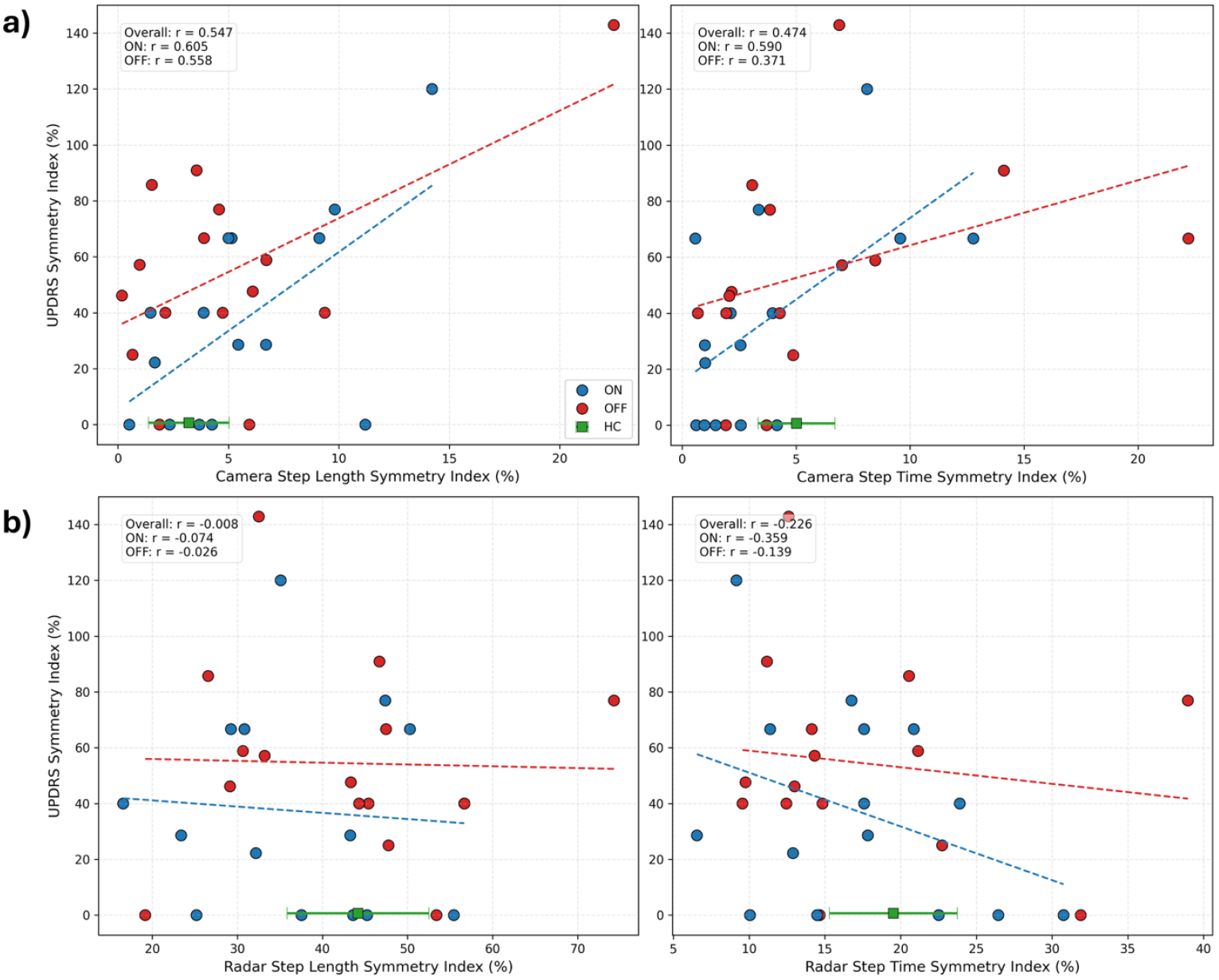
Gait symmetry correlations between clinical and sensor-derived measures. **(a)** Correlations between UPDRS III gait symmetry scores and camera-based step-length symmetry (left) and step-time symmetry (right). **(b)** Equivalent correlations for radar-based indices. Each point represents a Parkinson’s participant in the ON (blue) or OFF (red) state. Sensor-derived gait symmetry indices for the healthy control group are presented as green error bars at the bottom of each panel.

## Discussion

This study validates the reliability and demonstrates the usability of nearables, passive motion-sensing devices, for tracking gait patterns in PD. We demonstrate the sensitivity to differentiate people with mild PD from healthy older adults and within-patient sensitivity to changes in gait over their daily medication cycle. Prior gait studies in PD have relied on structured laboratory protocols or wearable sensors that require active participation. Our findings demonstrate that passive, non-contact sensing can reliably detect disease-related and medication-related gait alterations in an ecologically valid environment, without burdening the participant. Additionally, the study reports step-level asymmetric measures, with a notable association observed in camera-derived metrics. Overall, these findings exhibit the feasibility of using passive sensors to capture clinically meaningful motor changes in PD.

While there is an increase in the use of wearable devices in Parkinson’s patients, including recent guidelines from the UK’s National Institute for Health and Care Excellence (NICE)(40), those devices can burden patients. Recent research addresses the practical challenges of deploying wearable sensor systems for long-term, continuous monitoring in real-world environments(20). Although feasible for monitoring mobility and medication adherence, technical and usability issues reduced data completeness and satisfaction (41). Older participants reported greater difficulty using the system. Similar trends have been reported in other sensor-based studies, where robustness, validation in unsupervised settings, and accessibility remain major barriers(42–44).

Passive sensing technologies show promising potential for addressing these limitations. Our study demonstrates consistent within-device and between-device performance despite inherent modality-specific variability. When comparing camera and radar measurements, the camera tended to overestimate distances; this might be attributed to inherent differences in how each sensor tracks limb segments compared to the overall center of mass. Nevertheless, this overestimation was consistent; thus, though above the identity line, the correlation between devices was high. Moreover, both camera and radar demonstrated high sensitivity in discriminating PD from HC based on stride length metrics. Stride length impairment is a well-established and significant marker of motor deficits in PD. Multiple studies reported stride length reductions with disease progression, while stride frequency tends to remain relatively stable or variably affected(1,45,46). Dysfunction in basal ganglia–cortical circuits disrupts movement amplitude, as reflected in improvements in gait symptoms and parameters when these pathways are targeted with dopaminergic therapy(47–50). In our results, radar exhibited the highest sensitivity in detecting both group-level differences between PD and HC and within-participant differences between PD medication states. A similar but weaker trend was observed in gait speed.

Although mild gait asymmetry occurs in healthy individuals, temporal asymmetries increase in early PD, and step-level metrics provide a more reliable assessment of symmetry as they directly capture interlimb coordination(51–53). Increased gait asymmetry is considered a key contributor to motor instability in PD and may amplify symptoms such as balance loss, especially when not considered during interventions like deep brain stimulation (DBS)(54,55). For instance, stimulation of the subthalamic nucleus (STN) has been reported to induce increased step time asymmetry and dyscoordination in some patients(56). The symmetry index in this study quantified the magnitude of asymmetry, revealing a moderately strong correlation between camera-derived step length asymmetry and the UPDRS motor asymmetry index. This association suggests that the camera-based metrics captured clinically meaningful limb imbalance, consistent with established lateralized motor dysfunction in PD. Our radar asymmetry metric failed to capture it here due to the limitation in capturing individual limbs with a single radar, as described in our previous work(36). Future work will integrate multiple sensors or receivers into a scalable system to enable limb-specific sensing, enhance asymmetry detection, and capture more gait features.

While providing within-patient evidence for device sensitivity to gait fluctuations, the statistical power might have been limited by the sample size, and the generalizability to advanced PD might be limited by the mild symptoms of our participants. Yet, the main limitation of this study is that while it was performed in a home-like environment, it was not in the home, and the walk was instructed. Future studies should replicate this finding in patients’ homes while they walk around the house without being instructed. In addition, further validation, including comparisons to common wearable systems, would strengthen the validity and reproducibility of the passive sensing approach. Finally, the short walking distance (4m) constrains the assessment of continuous gait variability and turning dynamics, which should be addressed in future activities of daily living.

## Conclusion

By capturing disease- and medication-related changes passively, radar and camera systems could complement clinic-based UPDRS assessments to provide objective, longitudinal markers of motor symptoms and their fluctuations. While camera-derived metrics capture spatial asymmetry linked to UPDRS-rated motor lateralization, radar offers enhanced sensitivity to dopaminergic state changes. Together, these passive sensing modalities provide a foundation for continuous, contact-free motor assessment, advancing the development of digital biomarkers for PD management in the home.

## Data Availability

Data produced in the present study are available upon reasonable request to the authors.

## Acknowledgement

This work was supported by the UK Dementia Research Institute Care Research & Technology Centre (UK DRI CR&T). CH was supported by the Engineering and Physical Sciences Research Council (EPSRC) Doctoral Training Partnership (DTP). We thank all the participants for their time and commitment.

## Authors’ Roles

**Juyoung Jenna Yun**: Methodology, Data Curation, Formal analysis, Writing – Original Draft & Review & Editing, Visualization; **Charalambos Hadjipanayi**: Methodology, Data Curation, Formal analysis, Writing – Review & Editing; **Arya Jahangiri**: Methodology, Formal analysis, Writing – Review & Editing; **Alan Bannon**: Data Curation, Writing – Review & Editing; ; **Tim Constandinou**: Validation, Writing – Review & Editing; **Shlomi Haar**: Conceptualization, Methodology, Validation, Data Curation, Writing – Original Draft, Writing – Review & Editing, Supervision;

## Notes

### Competing Interest Statement

The authors have declared no competing interest.

### Funding Statement

This study did not receive any funding.

### Author Declarations

The study was approved by the Imperial College Research Ethics Committee (ICREC) and conducted in accordance with the Declaration of Helsinki.

